# A genome-wide association study of total child psychiatric problems scores

**DOI:** 10.1101/2020.06.04.20121061

**Authors:** Alexander Neumann, Ilja M. Nolte, Irene Pappa, Tarunveer S. Ahluwalia, Erik Pettersson, Alina Rodriguez, Andrew Whitehouse, Catharina E.M. van Beijsterveldt, Beben Benyamin, Anke R. Hammerschlag, Quinta Helmer, Ville Karhunen, Eva Krapohl, Yi Lu, Peter J. van der Most, Teemu Palviainen, Beate St Pourcain, Ilkka Seppälä, Anna Suarez, Natalia Vilor-Tejedor, Carla M. T. Tiesler, Carol Wang, Amanda Wills, Ang Zhou, EAGLE behavior & cognition group, Silvia Alemany, Hans Bisgaard, Klaus Bønnelykke, Gareth E. Davies, Christian Hakulinen, Anjali K. Henders, Elina Hyppönen, Jakob Stokholm, Meike Bartels, Jouke-Jan Hottenga, Joachim Heinrich, John Hewitt, Liisa Keltikangas-Järvinen, Tellervo Korhonen, Jaakko Kaprio, Jari Lahti, Marius Lahti-Pulkkinen, Terho Lehtimäki, Christel M. Middeldorp, Jackob M. Najman, Craig Pennell, Chris Power, Albertine J. Oldehinkel, Robert Plomin, Katri Räikkönen, Olli T Raitakari, Kaili Rimfeld, Lærke Sass, Harold Snieder, Marie Standl, Jordi Sunyer, Gail M. Williams, Marian J. Bakermans-Kranenburg, Dorret I. Boomsma, Marinus H. van IJzendoorn, Catharina A. Hartman, Henning Tiemeier

**Affiliations:** Department of Child and Adolescent Psychiatry/Psychology, Erasmus University Medical Center Rotterdam, the Netherlands; Lady Davis Institute for Medical Research, Jewish General Hospital, Montreal, QC, Canada; Department of Epidemiology, University of Groningen, University Medical Center Groningen, Groningen, The Netherlands; COPSAC, Copenhagen Prospective Studies on Asthma in Childhood, Herlev and Gentofte Hospital, University of Copenhagen, Copenhagen, Denmark; Steno Diabetes Center Copenhagen, Gentofte, Denmark; Department of Medical Epidemiology and Biostatistics, Karolinska Institutet, Stockholm, Sweden; Department of Epidemiology and Biostatistics, School of Public Health, Imperial College London, London, UK; Telethon Kids Institute, University of Western Australia, Perth, Australia; Netherlands Twin Register, Dept Biological Psychology, Vrije Universiteit, Amsterdam, The Netherlands; Australian Centre for Precision Health, University of South Australia Cancer Research Institute, School of Health Sciences, University of South Australia, Adelaide, Australia; South Australian Health and Medical Research Institute, Adelaide, Australia; Child Health Research Centre, University of Queensland, Brisbane QLD, Australia; Biological Psychology, Vrije Universiteit Amsterdam, Amsterdam, the Netherlands; Amsterdam Public Health research institute, Amsterdam, the Netherlands; Centre for Life-Course Health Research, University of Oulu, Oulu, Finland; Social, Genetic, and Developmental Psychiatry Centre, Institute of Psychiatry, Psychology and Neuroscience, King’s College London, London, United Kingdom; Institute for Molecular Medicine Finland (FIMM), University of Helsinki, Helsinki, Finland; MRC Integrative Epidemiology Unit, Department of Population Health Sciences, Bristol Medical School, University of Bristol, Bristol BS8 2BN, UK; Language and Genetics Department, Max Planck Institute for Psycholinguistics, 6525 XD Nijmegen, The Netherland; Donders Institute for Brain, Cognition and Behaviour, Radboud University, 6525 EN Nijmegen, The Netherlands; Department of Clinical Chemistry, Fimlab Laboratories, Tampere 33520, Finland; Department of Clinical Chemistry, Finnish Cardiovascular Research Center - Tampere, Faculty of Medicine and Health Technology, Tampere University, Tampere 33014, Finland; Department of Psychology and Logopedics, Faculty of Medicine, University of Helsinki; Center for Genomic Regulation (CRG), the Barcelona Institute of Science and Technology, Barcelona, Spain; BarcelonaBeta Brain Research Center (BBRC) – Pasqual Maragall Foundation, Barcelona, Spain; Department of Clinical Genetics, Erasmus Medical Center, Rotterdam, the Netherlands; Universitat Pompeu Fabra (UPF), Barcelona, Spain; Institute of Epidemiology, Helmholtz Zentrum München - German Research Center for Environmental Health, Neuherberg, Germany; LMU – Ludwig-Maximilians-Universität Munich, Div. Metabolic and Nutritional Medicine, Dr. von Hauner Children’s Hospital, University of Munich Medical Center, Munich, Germany; School of Medicine and Public Health, Faculty of Medicine and Health, The University of Newcastle, Newcastle, New South Wales, Australia; Division of Substance Dependence, Department of Psychiatry, University of Colorado Anschutz Medical Campus; Institute for Behavioral Genetics, University of Colorado Boulder; ISGlobal, Centre for Research in Environmental Epidemiology (CREAL), Barcelona, Spain; CIBER Epidemiology and Public Health (CIBERESP), Barcelona, Spain; Avera Institute for Human Genetics, Sioux Falls, South Dakota, USA; Institute of Molecular Bioscience, University of Queensland, Brisbane QLD, Australia; Population, Policy and Practice, UCL Great Ormond Street Institute of Child Health, London, UK; Department of Pediatrics, Naestved Hospital, Naestved, Denmark; Institute and Clinic for Occupational, Social and Environmental Medicine, University Hospital, LMU Munich, Munich, Germany; Department of Psychology and Neuroscience, University of Colorado Boulder; Department of Public Health, University of Helsinki, Helsinki, Finland; Child and Youth Mental Health Service, Children’s Health Queensland Hospital and Health Service, Brisbane QLD, Australia; Public Health, University of Queensland, Brisbane QLD, Australia; Interdisciplinary Center Psychopathology and Emotion Regulation (IPCE), University of Groningen, University Medical Center Groningen, Groningen, The Netherlands; Department of Clinical Physiology and Nuclear Medicine, Turku University Hospital, Turku 20521, Finland; Research Centre of Applied and Preventive Cardiovascular Medicine, University of Turku, Turku 20014, Finland; Department of Clinical Physiology and Nuclear Medicine, Turku University Hospital, Turku, Finland; IMIM (Hospital del Mar Medical Research Institute), Barcelona, Spain; Clinical Child & Family Studies, Vrije Universiteit Amsterdam, the Netherlands; School of Clinical Medicine, University of Cambridge, UK; Department of Psychiatry, University of Groningen, University Medical Center Groningen, Groningen, The Netherlands; Department of Social and Behavioral Science, Harvard TH Chan School of Public Health, Boston, MA, USA

**Author notes:** **CORRESPONDING AUTHOR** Henning Tiemeier, Erasmus University Medical Center Rotterdam, Department of Child and Adolescent Psychiatry/Psychology, Room KP-2822, PO-Box 2060, 3000 CB Rotterdam, The Netherlands.

## Abstract

Substantial genetic correlations have been reported across psychiatric disorders and numerous cross-disorder genetic variants have been detected. To identify the genetic variants underlying general psychopathology in childhood, we performed a genome-wide association study using a total psychiatric problem score. We analyzed 6,844,199 common SNPs in 38,418 school-aged children from 20 population-based cohorts participating in the EArly Genetics and Lifecourse Epidemiology (EAGLE) consortium. The SNP heritability of total psychiatric problems was 5.4% (SE=0.01) and two loci reached genome-wide significance: rs10767094 and rs202005905. We also observed an association of *SBF2*, a gene associated with neuroticism in previous GWAS, with total psychiatric problems. The genetic effects underlying the total psychiatric problem score were shared with known genetic variants for common psychiatric disorders only (attention-deficit/hyperactivity disorder, anxiety, depression, insomnia) (r_G_ > 0.49), but not with autism or the less common adult disorders (schizophrenia, bipolar disorder, or eating disorders) (r_G_ < 0.01). Importantly, the total psychiatric problem score also showed at least a moderate genetic correlation of with intelligence, educational attainment, wellbeing, smoking, and body fat (r_G_ > 0.29).The results suggest that many common genetic variants are associated with childhood psychiatric symptoms and related phenotypes in general instead of with specific symptoms. Further research is needed to establish causality and pleiotropic mechanisms between psychiatric disorders and related traits.

## Introduction

Psychiatric disorders are moderately heritable, on average about 30-50% of the variability in symptoms can be explained by genetic differences between individuals. [1] The joint effect of common single nucleotide polymorphisms (SNP heritability) explains 5% to 30% of the variance in psychiatric disorders in adults.[2] Similar levels have been reported for behavioral and emotional symptoms in children, although there is large variability depending on child age and informant.[3, 4] A focus on childhood problems is particularly important, as many adult disorders can be traced back to problems in childhood.[5]

Recent family and molecular genetic studies demonstrated that much of the genetic effects underlying psychiatric disorders are not unique to particular diagnoses, but rather shared across several psychiatric diagnoses and symptoms.[2, 6–10] This phenomenon is known as cross-phenotype association and suggests pleiotropy, i.e. the influence of a genetic variant on multiple traits,[11] and may be an explanation for the extensive co-occurrence of mental disorders.[12] Several lines of evidence support this notion. First, the SNP based genetic correlations between disorders from different domains, such as major depression, attention-deficit/hyperactivity disorder (ADHD), bipolar disorder and schizophrenia are moderate to high,[2] averaging 0.41[9]. Second, measures of global psychopathology in children showed a common SNP heritability between 16% and 38%.[8, 13] Third, a genome-wide association meta-analysis (GWAS) of eight psychiatric disorders (ADHD, anorexia, autism, bipolar, depression, obsessive compulsive disorder, schizophrenia and Tourette’s) identified 23 loci associated with at least four of these disorders.[14]

GWAS derived polygenic risk scores (PRS) for single disorders are good predictors of general psychopathology. For instance, a PRS for ADHD was more strongly associated with a general psychopathology factor than with specific hyperactivity or attention problems adjusted for general psychopathology.[15] In another study a composite PRS based on eight GWAS was associated with general psychopathology in childhood.[16] These cross-phenotype associations present a challenge in interpreting GWAS results that typically target a single disorder, raising the question of whether a multi-disorder approach would be more informative.

Previous GWAS of childhood disorders, such as autism spectrum disorders, ADHD, aggression and internalizing disorders,[4, 17–19] have provided insights into the genetic architecture of child psychiatric problems and into the genetic correlations between childhood psychiatric problems. However, with notable exceptions of a large recent ADHD study[20] and a GWAS on autism spectrum disorder[17], these studies mostly failed to identify individual genome-wide significant loci. Besides increasing the sample size, some researchers propose the inclusion of related phenotypes in analyses to increase power. [21, 22] Genetic loci with pleiotropic effects may be missed in a GWAS of single psychiatric disorders. If a variant only modestly increases the risk of symptoms from different domains, any association with a specific disorder may be too weak to be detected. A focus on global psychopathology increases the power to detect unspecific genetic loci, which are associated with global psychiatric vulnerabilities. A previous GWAS[14] examined multiple disorder simultaneously, but analyses of multiple dimensional measures of psychiatric problems in childhood are lacking. This approach is arguably particularly promising in childhood given the less clearly expressed symptoms and the low homotypic but high heterotypic stability of problems,[23] i.e. the changing of symptoms from one domain to another.

Our aim was to identify genetic loci associated with a total psychiatric problem score representing a variety of psychiatric problems including internalizing, externalizing, attention, neurodevelopmental and other psychiatric problems. To identify these genetic variants, we performed a GWAS meta-analysis within the EArly Genetics and Lifecourse Epidemiology (EAGLE) consortium (https://www.eagle-consortium.org/). Finally, we estimated genetic correlations of the total psychiatric problem score with various single child and adult psychiatric, psychological, neurological and lifestyle or educational characteristics.

## Materials and Methods

### Participants

Cohorts from the EAGLE consortium with parent-rated measures of psychiatric symptoms in the age range 5-16 years were invited to participate in the project Twenty cohorts from Europe, the US and Australia contributed data to this meta-analysis. See Table 1 and supplementary materials for cohort descriptions. Parents provided informed consent and study protocols were approved by local ethics committees. We restricted the analysis to children of European ancestry to avoid population stratification bias. In total data from 38,418 participants with a mean age of 9.9 years (SD=2.02) were meta-analyzed. This study was originally planned with a discovery-replication design. However, the obtained sample-size was not sufficiently large to split the sample, and we opted for maximizing power in discovery analyses.

**Table 1:** Phenotype Characteristics

| Cohort | n | Instrument | Domains | Informant | Age years | Age SD | Score Mean | Score SD | % Female |
| --- | --- | --- | --- | --- | --- | --- | --- | --- | --- |
| 1958BC-T1DGC | 2170 | Rutter | Int,Ext | Maternal | 11.3 | 0.1 | 6.2 | 3.4 | 51 |
| 1958BC-WTCCC | 2261 | Rutter | Int,Ext | Maternal | 11.3 | 0.2 | 6.2 | 3.4 | 48 |
| ALSPAC | 5461 | SDQ | Int,Ext | Maternal | 9.6 | 0.1 | 6.7 | 4.8 | 49 |
| BREATHE | 1618 | SDQ | Int,Ext | Both | 8.3 | 3.9 | 8.1 | 5.1 | 48 |
| CADD | 358 | CBCL 4-18 | Int,Ext,Sleep,TP,EP,PDD | Both | 13.0 | 2.6 | 16.2 | 21.9 | 28 |
| CATSS | 6498 | A-TAC | Int,Ext,EP,PDD | Both | 12.0 | 0.0 | 5.4 | 7.5 | 49 |
| COPSAC2010 | 547 | SDQ 4-10 | Int,Ext | Both | 6.0 | 0.3 | 7.1 | 4.7 | 48 |
| FinnTwin12 | 959 | MPNI | Int,Ext | Both | 11.4 | 0.3 | 11.3 | 6.8 | 53 |
| GenR | 1847 | CBCL 6-18 | Int,Ext,Sleep,TP,EP,PDD | Maternal | 9.7 | 0.3 | 17.3 | 15.2 | 51 |
| Gini-Lisa | 1389 | SDQ | Int,Ext | Maternal | 10.0 | 0.2 | 7.3 | 5.2 | 48 |
| Glaku | 312 | CBCL 6-18 | Int,Ext,Sleep,TP,EP,PDD | Maternal | 12.1 | 1.0 | 21.7 | 16.8 | 52 |
| INMA | 745 | SDQ | Int,Ext | Both | 5.1 | 0.8 | 8.9 | 5.0 | 38 |
| MUSP | 1156 | CBCL 6-18 | Int,Ext,Sleep,TP,EP,PDD | Maternal | 13.9 | 0.3 | 30.5 | 19.8 | 61 |
| NFBC1986 | 3346 | Rutter | Int,Ext | Maternal | 7.8 | 0.2 | 2.6 | 2.1 | 51 |
| NTR I | 2563 | CBCL 6-18 | Int,Ext,Sleep,TP,EP,PDD | Maternal | 9.9 | 1.0 | 19.3 | 15.9 | 52 |
| NTR II | 2960 | CBCL 6-18 | Int,Ext,Sleep,TP,EP,PDD | Maternal | 9.6 | 1.0 | 19.1 | 16.6 | 53 |
| RAINE | 1366 | CBCL 4-18 | Int,Ext,Sleep,TP,EP,PDD | Both | 10.6 | 0.2 | 21.1 | 18.6 | 48 |
| TCHAD | 2111 | CBCL 6-18 | Int,Ext,Sleep,TP,EP,PDD | Both | 13.0 | 0.0 | 11.7 | 12.5 | 51 |
| TEDS | 2707 | SDQ | Int,Ext | Both | 11.3 | 0.7 | 7.0 | 5.0 | 54 |
| TRAILS | 1283 | CBCL 6-18 | Int,Ext,Sleep,TP,EP,PDD | Maternal | 11.1 | 0.6 | 0.2 | 0.2 | 52 |
| YFS | 1352 | HES | Int,Ext | Maternal | 10.6 | 3.3 | 14.7 | 6.8 | 54 |

**n** sample size
**Domains** covered by instrument: Internalizing (Int), Externalizing (Ext), Sleep, Thought Problems (TP), Eating Problems (EP), Pervasive Developmental Disorder Score (PDD)
**Informant** questionnaire filled in by only mothers (maternal) or by either father or mother (both)
**SD** standard deviation

### Outcome

Psychiatric problems were assessed with parent-rated questionnaires at the assessment wave closest to age 10 years. All items of a broad psychiatric questionnaire were summed into a single total psychiatric sum score. In all cohorts internalizing, externalizing and attention problems were assessed in some questionnaires items on sleep, thought, eating problems, and pervasive developmental disorders were included in the total problem score (Table 1). Instruments included the Child Behavior Checklist (CBCL)[24], Strengths and Difficulties Questionnaire (SDQ)[25], parental version of the Multidimensional Peer Nomination Inventory (MPNI)[26], Rutter Children’ Behaviour Questionnaire[27], the Autism–Tics, AD/HD and other Comorbidities inventory (A-TAC)[28], and items derived from the Health Examination Survey[29].

We applied a log transformation plus 1 to avoid bias due to non-normal residuals and influential observations. Because different scales were used, the log-transformed scores were converted to a z-score within cohorts to make units comparable across cohorts.

### Genotyping and QC

Genotyping was performed using genome-widearrays. Cohort-specific pre-imputation quality control (QC) was performed using established protocols. In all cohorts, SNPs were imputed to the 1000 Genomes Phase 1 or Phase 3 reference panel.[30] Each cohort performed a GWAS and summary results were collected for meta-analysis. We omitted the X-chromosome from further analysis as most cohorts had no information available on X-linked SNPs. Pre-meta-analysis QC was performed with EasyQC and QCGWAS.[31–33] The QC steps are summarized in Figure S1. After meta-analysis, we excluded SNPs with low minor allele frequency (MAF < 5%), sample size (<5000), or with data from a small number of cohorts (<5). Finally, we checked the pooled results for spurious inflation by examining QQ-plots of the p-value distribution and by examining the LD score regression intercept (see statistical analysis). Full genetic methods and quality control per cohort can be found in Table S1 and Table S2.

### Statistical analysis

#### Single SNP associations and meta-analysis

The z-scores of the total psychiatric problems scores were related to the SNP dosages in a linear model. Covariates included gender, age at assessment and principal components of ancestry. The number of dimensions (1-10) were specified by each cohort.. CATSS and TCHAD additionally used a random effect to account for familial relatedness. FinnTwin12 and NTR applied a mixed model with two random effects to control for population stratification and relatedness. We pooled the results from the individual cohorts using an inverse-variance weighted fixed-effects meta-analysis. R 3.4.3 was used for QC, data preparation and analysis of results.[34] Meta-soft 2.0.1 was used for the meta-analysis of single SNP associations.[35] [35] The individual cohort results after quality control were examined and meta-analyzed independently by the first and second author with consistent results. Genome-wide significance was set at p<5E-08.

We also used the FUMA web tool[36] to explore potential functional implications of any identified variants. We reviewed positional mapping, eQTL analyses and chromatin interactions with all available databases (date: 2019-06-30). We also performed a lookup in the mQTL[37] database, to check for potential influences on gene expression via DNA methylation.

#### Gene-based and expression analysis

We performed gene-based tests using MAGMA[38] in FUMA. MAGMA estimates the joint effect of all SNPs within a gene, while accounting for the LD structure and gene size. We tested 18,168 protein coding genes and thus the p-value significance threshold was set at 3e-6 based on Bonferroni correction.

Second, we tested, whether the results from the gene-based tests were related to gene expression in several tissues. Specifically, we used MAGMA to test whether the strength of association between genes and the total psychiatric problem score was related to the mean gene expression level in a specific tissue, while considering average expression levels. Given that we expected gene variants to act via brain pathways, we tested expression in 13 brain regions (Table S3). However, as gene effects may impact the brain indirectly via other tissues, we also investigated gene expression levels on an organ level (Table S4). Gene expression levels were obtained from the GTEx 7 database.[39]

Third, we further examined whether the predicted gene expression of selected genes was related to total psychiatric problems. We selected genes, that were (functionally) annotated to genome-wide hits, or that were genome-wide significant according to gene-based tests. To correlate gene expression with total psychiatric problems, we used a transciptome-wide association study (TWAS) approach.[40] In short, gene expression in a tissue is imputed based on expression information from the GTEx 7 database for a specific tissue and then correlated with a phenotype, as inferred from GWAS summary statics. We chose to examine expression in the basal ganglia post-hoc, as genes most strongly associated with total psychiatric problems tended to be expressed in this brain region. We also performed a lookup on TWAS hub, to examine whether gene expression by a gene identified in this study has previously been associated with other phenotypes.[41, 42]

#### SNP heritability and genetic correlations

We estimated the SNP heritability of total psychiatric problem scores with LD score regression.[43] We used the online tool LD Hub[44] to estimate common SNP heritability and genetic correlations with various psychiatric, psychological, neurological and lifestyle or educational characteristics. To compute the genetic correlations we selected published GWAS summary statistics available on LD Hub, except genetic correlations with anxiety symptoms[45], which were computed locally with ldsc 1.0.0.

## Results

### Spurious inflation and SNP Heritability

We tested 6,844,199 SNPs after quality control. The QQ-plot (Figure 1) showed some inflation, however, the LD score intercepts was close to 1 (β_0_ = 1.01, SE=0.01), suggesting that the inflation was due to a true signal rather than spurious associations. The SNP heritability was estimated at 5.4% (SE = 0.013).

**Figure 1:**
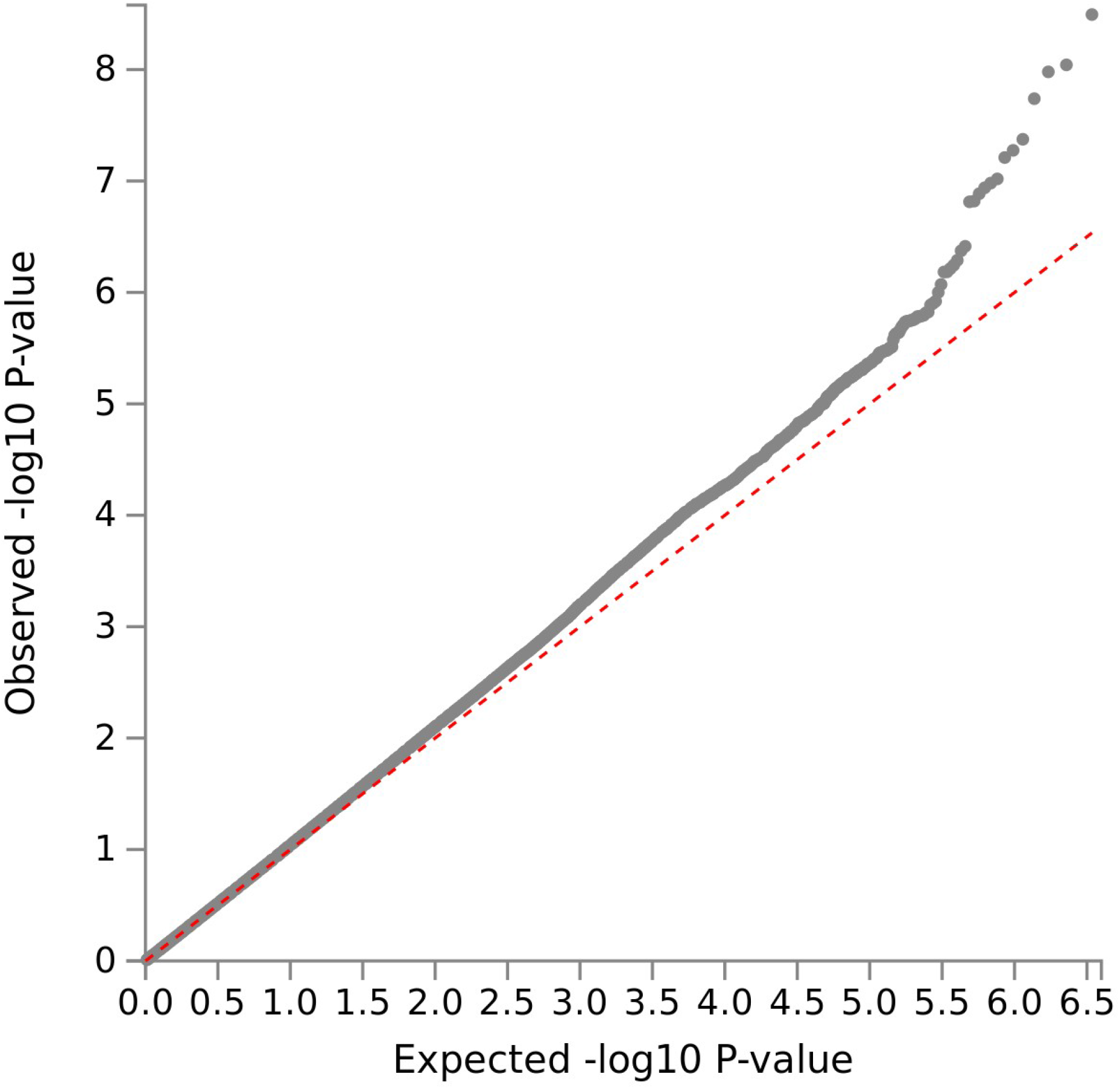
Quantile-quantile plot of observed -log 10 p values vs expected -log 10 p values assuming chance findings in single SNP analysis. Diagonal line indicates a p value distribution compatible with chance finding. Upward deviations indicate p values more significant than expected.

### SNP based tests

Two loci on chromosome 11 were genome-wide significant, see Figure 2. One locus is located around lead SNP rs10767094, which showed an increase of 0.08SD in total psychiatric problems per A allele (SE=0.01, p=3E-09, n=8,216) (Figure S2). The A allele is very common with an average frequency of 48% across the cohorts, but the SNP’s average imputation quality was a moderate 50% (Info/R^2^). Information on this locus was only available in 27% of participants (8 cohorts). The SNP showed a moderate amount of effect heterogeneity (I^2^=47.6%). Also on chromosome 11 an insertion/deletion variant (InDel) was genome-wide significant. A deletion of the A allele at rs202005905 was associated with an increase of 0.08SD in total psychiatric problems (SE=0.01, p=4E-08, n=15,886, Figure S3). Deletion prevalence was on average 16%, but again the imputation quality was modest with 52%, information was available in 41% of participants (9 cohorts) and the genetic variant showed moderate effect heterogeneity (I^2^=59.6%).

**Figure 2:**
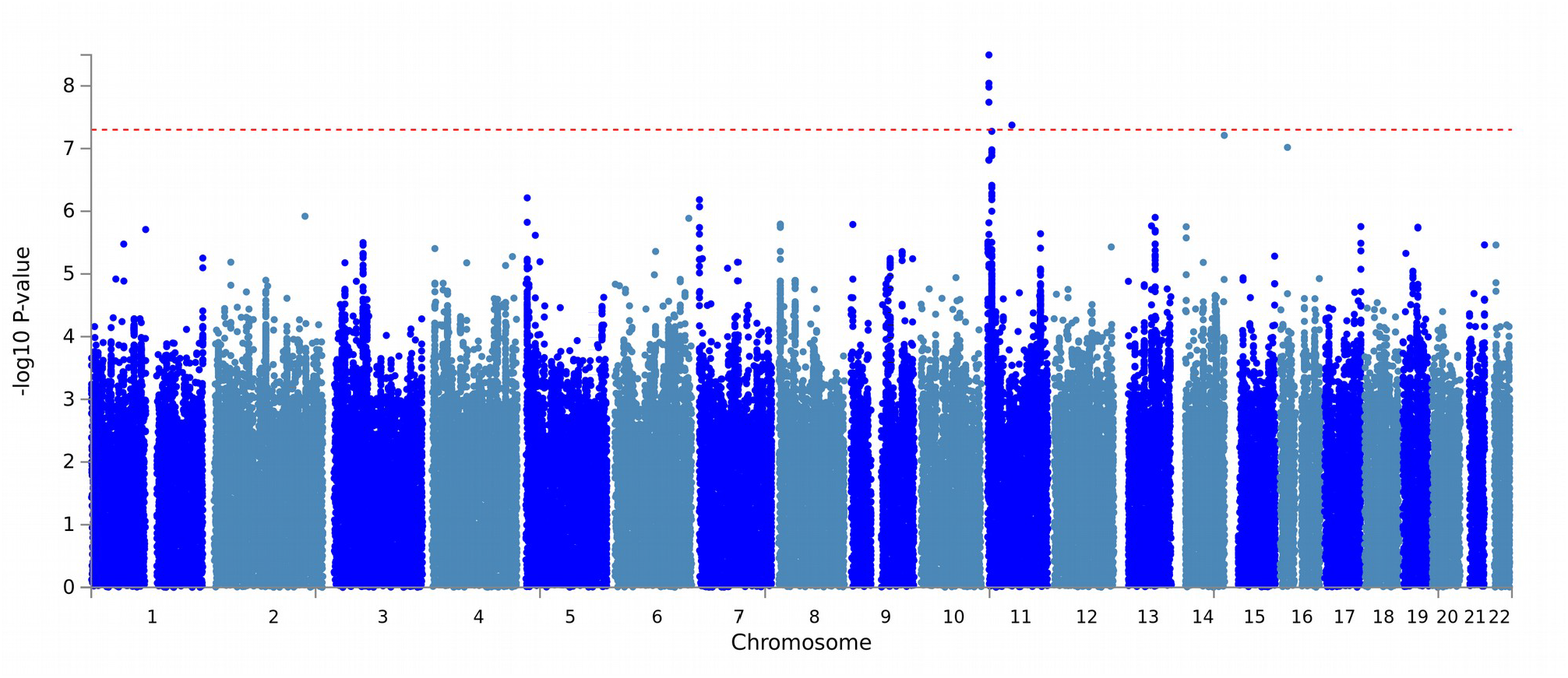
Manhattan plot of -log 10 p values vs SNP position for single SNP analysis. SNPs above the red horizontal line indicate genome-wide significant findings.

The SNP rs10767094 lies in the intron of an uncharacterized gene and rs202005905 lies in an intergenic region with no nearby genes. A FUMA eQTL and chromatin interaction analysis did not reveal any interactions with genes. The mQTL database did not list any associations with DNA methylation.

The third top locus did not reach genome-wide significance, but is of interest for its location in a gene previously implicated in neuroticism[46, 47] as well as being very close to genome-wide significance. The SNP rs72854494 lies within the gene *SBF2*. The T allele was associated with 0.05SD lower total psychiatric problems (SE=0.01, p=5E-08, n=38,330)(Figure S4). This association showed no heterogeneity (I^2^=0.0%) among the cohorts. The T allele occurred on average in 14% across cohorts, with a very good imputation quality of 96%. FUMA eQTL and chromatin interaction analysis, as well as a lookup in mQTL DB did not reveal any further information on functional association. Results for all SNPs with genome-wide suggestive p-values (p<5E-06) can be found in Table 2.

**Table 2:** SNPs with genome-wide significant (p<5E-08) and suggestive (p<5E-07) results

| SNP | Chr | BP | EA | OA | EAF | n<sub>stu</sub> | n | $\beta$ | SE | p | I<sup>2</sup> |
| --- | --- | --- | --- | --- | --- | --- | --- | --- | --- | --- | --- |
| rs10767094 | 11 | 3477509 | A | G | 0.48 | 6 | 8216 | 0.08 | 0.01 | 3E-09 | 47.6 |
| rs12098951 | 11 | 3478953 | A | G | 0.48 | 8 | 10417 | 0.08 | 0.01 | 9E-09 | 52.3 |
| rs10767093 | 11 | 3477421 | T | A | 0.48 | 8 | 10408 | 0.08 | 0.01 | 1E-08 | 40.3 |
| rs10767096 | 11 | 3477891 | T | C | 0.53 | 8 | 10382 | 0.07 | 0.01 | 2E-08 | 47.6 |
| rs202005905 | 11 | 54733705 | I | D | 0.84 | 9 | 15886 | -0.08 | 0.01 | 4E-08 | 59.6 |
| rs72854494 | 11 | 9946312 | T | C | 0.86 | 21 | 38330 | -0.05 | 0.01 | 5E-08 | 0.0 |
| rs188216744 | 14 | 106478354 | T | C | 0.55 | 5 | 7045 | 0.08 | 0.01 | 6E-08 | 87.2 |
| rs115749482 | 16 | 16754648 | A | G | 0.81 | 5 | 6930 | 0.08 | 0.01 | 1E-07 | 85.6 |
| rs59076561 | 11 | 9951438 | G | T | 0.86 | 20 | 35645 | -0.05 | 0.01 | 1E-07 | 0.0 |
| rs113227893 | 11 | 9944120 | D | I | 0.86 | 18 | 33850 | -0.05 | 0.01 | 1E-07 | 0.2 |
| rs67456791 | 11 | 9944108 | G | A | 0.86 | 20 | 35533 | -0.05 | 0.01 | 1E-07 | 0.0 |
| rs10767095 | 11 | 3477568 | G | A | 0.48 | 8 | 10413 | 0.07 | 0.01 | 2E-07 | 43.8 |
| rs10834158 | 11 | 3477887 | A | G | 0.52 | 9 | 11682 | 0.07 | 0.01 | 2E-07 | 56.1 |
| rs116657155 | 11 | 9954242 | A | G | 0.85 | 20 | 35619 | -0.05 | 0.01 | 4E-07 | 0.0 |
| rs60713856 | 11 | 9955418 | G | C | 0.85 | 20 | 35579 | -0.05 | 0.01 | 4E-07 | 0.0 |
| rs140557414 | 11 | 9953387 | A | C | 0.85 | 20 | 35632 | -0.05 | 0.01 | 5E-07 | 0.0 |
| rs57331333 | 11 | 9956272 | T | C | 0.85 | 20 | 35570 | -0.05 | 0.01 | 6E-07 | 0.0 |
| rs34543113 | 5 | 3339568 | G | A | 0.70 | 20 | 35612 | -0.04 | 0.01 | 6E-07 | 0.0 |
| rs11042555 | 11 | 9957159 | T | C | 0.85 | 20 | 35571 | -0.05 | 0.01 | 7E-07 | 0.0 |
| rs36189439 | 7 | 323206 | A | G | 0.58 | 6 | 7814 | 0.07 | 0.01 | 7E-07 | 69.2 |

**Chr** Chromosome**BP** Basepair Position (Build 37 map)**EA** Effect Allele**OA** Other Allele**EAF** Effect Allele Frequency**n<sub>stu</sub>** Number of Studies
**n** Sample Size
**$\beta$** Beta
**SE** standard error
**p** p-value
**$I^2$** Effect heterogeneity

### Gene-based test

Next we tested the association of 18,290 protein coding genes with the child total psychiatric problem score None of the genes reached genome-wide significance (Table S3, Figure S5 and Figure S6). We also post-hoc looked up the association of *SBF2*. The aggregate of 1,508 SNPs in *SBF2* showed a nominal significance of p=0.0004 (n=35,736). The full summary results can be found as supplementary data.

### Gene expression

We performed a MAGMA tissue expression analysis in 13 specific brain tissues (Table S4). Genes more strongly associated with total psychiatric problems tended to express particularly in four subcortical structures: caudate, putamen, anterior cingulate cortex and amygdala. However, this associations were not significant after correction for multiple testing. In addition we analyzed tissue expression for 30 tissues on an organ level, see Table S5. None of the organs had statistically significant associations, however, expression in the brain showed the strongest association (p=0.06).

The top two genome-wide significant loci were not linked to a characterized gene, thus we decided to perform a TWAS analysis only for *SBF2*. We found that higher predicted levels of *SBF2* in the basal ganglia were related to higher scores of total psychiatric symptoms (Z=+2.33, p=0.02) based on the best linear unbiased predictions (BLUP) of a random variable representing 489 SNPs. A lookup in the TWAS Hub database revealed, that predicted levels of *SBF2* gene products associate most with following phenotypes: neuroticism, body fat measures, red blood cell count, nervous feelings and worrying (http://twas-hub.org/genes/SBF2/).

### Genetic correlation

Next we quantified the extent to which the genetic associations of child psychiatric problems scores were shared with other phenotypes. After adjustment for false discovery rate, insomnia, depressive symptoms, neuroticism, cigarettes smoked per day, body fat, body mass index, number of children, and age of smoking initiation all showed positive genetic correlations between 0.29 and 0.60 with the total psychiatric problem score (Table 3) based on the results of independent GWAS in adults. The highest correlation of total psychiatric problems was with ADHD, but this association did not survive multiple testing correction (r_G_=0.86, SE=0.39, p=0.03, q=0.06). Subjective wellbeing, childhood IQ, college completion, years of schooling, intelligence and age of smoking initiation showed significant negative correlations with the total psychiatric problem score, ranging from − 0.66 to −0.42. Of the psychiatric phenotypes tested, the less common psychiatric disorders like schizophrenia, bipolar disorder, autism spectrum disorder, and anorexia were not genetically correlated with the total psychiatric problem score (rG < 0.01).

**Table 3:**
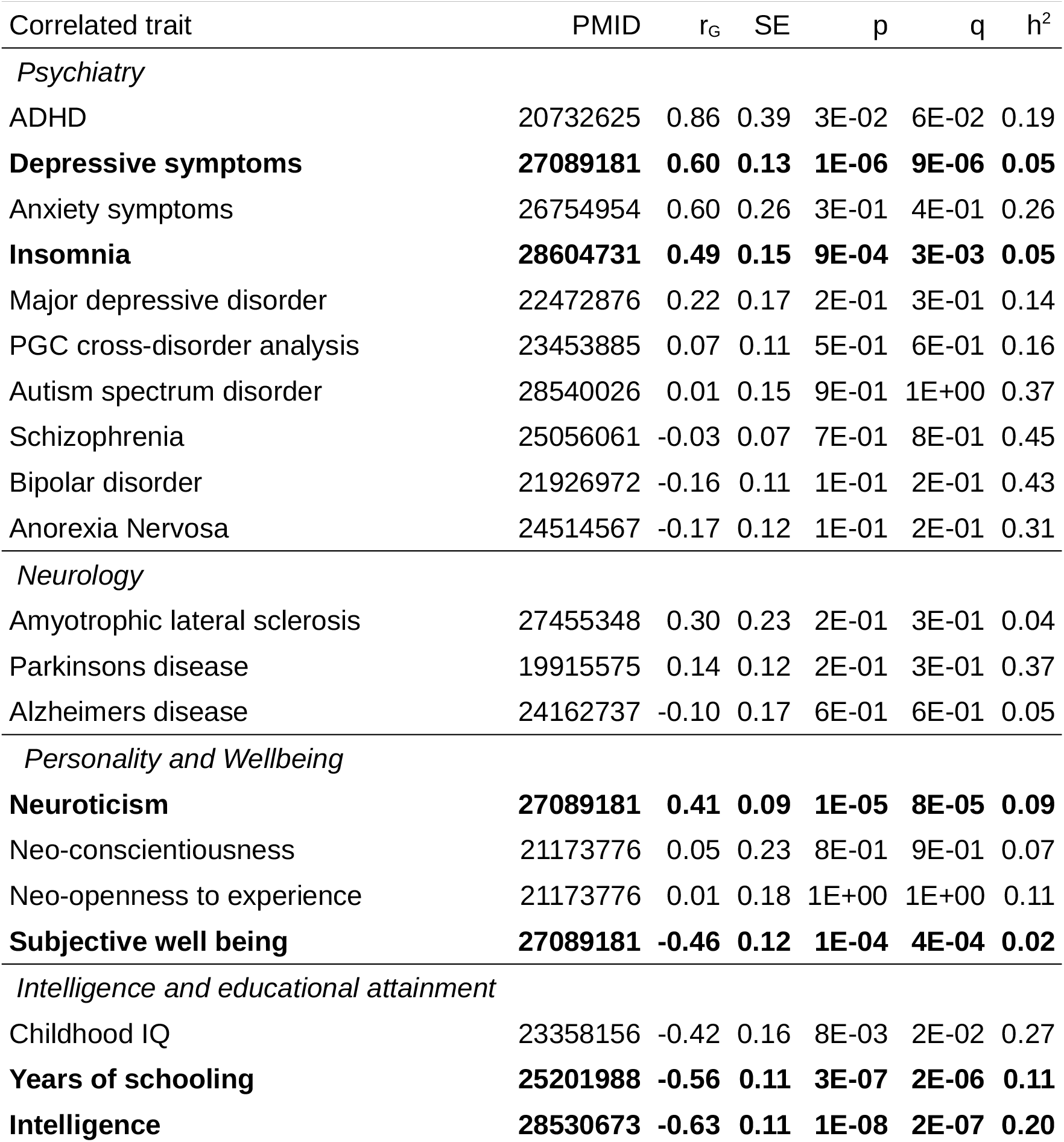
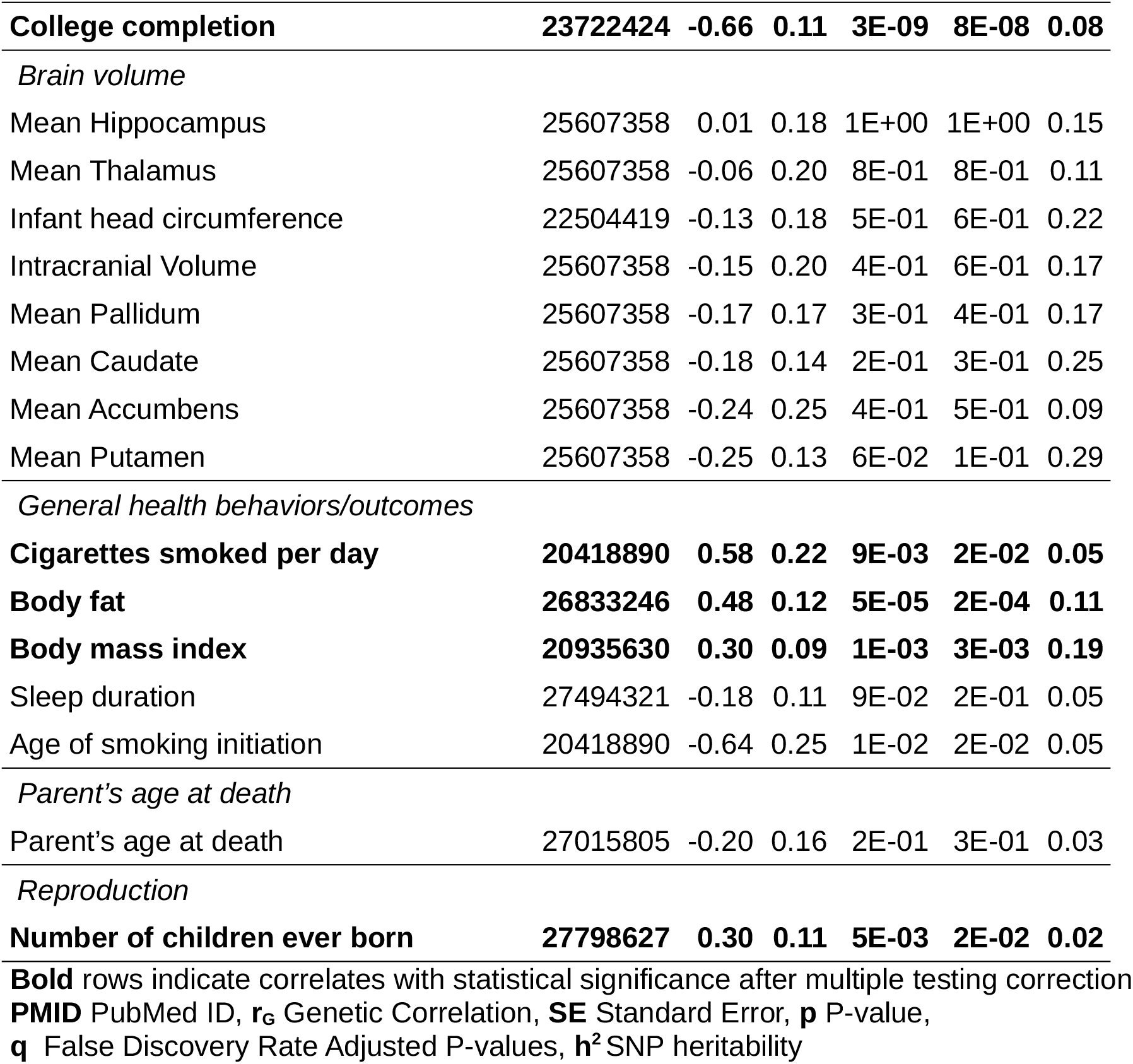
Genetic correlations based on LD score regression

## Discussion

The current study reports the first GWAS examining global psychopathology in children. Two genetic loci were genome-wide significant in the total sample. Additionally, we found support for the involvement of gene *SBF2* in the development of psychopathology. The genetic effects underlying global psychopathology were shared with common psychiatric disorders (ADHD, anxiety, depression, insomnia), but not with less common and on average more severe ones (schizophrenia, bipolar disorder, autism, eating disorders).

The two genome-wide significant variants are one SNP (rs10767094) and one InDel (rs202005905). To the best of our knowledge these variants have not been associated with psychiatric traits before. It is unclear, how exactly these variants or tagged causal variants may affect general psychopathology, as functional annotation for these loci is sparse. The modest imputation quality possibly affected study results as both variants failed quality control in most cohorts. Measurement error of the genotypes could explain the relatively high estimates heterogeneity.. An important next step would therefore be to replicate these SNPs using direct genotyping or denser arrays.

While just not genome-wide significant, the evidence for an involvement of *SBF2* with the lead SNP rs72854494 in total psychiatric problems is more convincing. This locus has been implicated in neuroticism based on two GWAS. In a GWAS of neuroticism[46] rs1557341, located in *SBF2*, showed genome-wide significance. In a second larger independent GWAS of 449,484 participants, *SBF2* showed a genome-wide significant effect for both neuroticism and worry in gene-based tests.[47] Furthermore, according to TWAS hub, the predicted gene products of *SBF2* correlate with neuroticism based on several GWAS. Neuroticism describes a disposition to experience negative emotions and a higher stress reactivity. It robustly and substantially associates with general psychopathology in children[8, 48], adolescence[49] and adults[50] (between r=0.13 and r=0.81). A twin study suggested that this correlation arises partly due to shared genetic causes[51] and in this GWAS the genetic correlations between total psychiatric problems and neuroticism were substantial as well, similar to the phenotypic association (r_G_=0.41). These results suggest that *SBF2* pleiotropically affects neuroticism and psychopathology, but the mechanisms would need to be explored further. Neuroticism has been hypothesized to contribute strongly to general psychopathology[52], thus it may mediate the effect of genetic variants on total psychiatric problems, but both phenotypes may also be independently affected. In regards to biology, human and mice studies points towards abnormal myelination as one of the consequences of *SBF2* alterations.[53, 54] We recently reported an association between lower global white matter integrity and higher levels of general psychopathology in school-aged children.[55] Thus, one may speculate that *SBF2* affects psychiatric problems via white matter development.

We additionally tested, whether genetic variants associated with total psychiatric problems were associated with gene expression in the brain. Association with gene expression in the limbic system of the brain showed the most support, but did not survive multiple testing correction. The findings are thus compatible with the possibility of a chance finding, but strong theoretical support for a major role of the limbic system exists. The limbic system includes evolutionary preserved regions responsible for emotion regulation and motivation[56], which were previously implicated in affective disorders, ADHD and OCD,[57, 58] and are a potential intervention target [59].

In this study we observed 5% SNP heritability, which is similar to the LD score estimated SNP heritability of continuously measured ADHD [4], depression[46] and anxiety symptoms[45] in population based cohorts. The total psychiatric problem scores were based on various instruments, which all included items for common psychiatric internalizing, attention, and externalizing symptoms. Therefore, it is not surprising that common psychiatric symptoms and disorders such as ADHD and depression shared 36% or more of the genetic variation with the total psychiatric problem score. The extent to which the questionnaires used in this study covered other less common problems, such as psychotic, bipolar or autistic symptoms varied greatly by instrument. Furthermore, age of onset for schizophrenia and bipolar disorder is typically in late adolescence and early adulthood.[60–62] For autism spectrum disorder, the age of onset is early, but the prevalence in the cohorts was low. Thus the total psychiatric problem score covered broad symptomatology but was not representative of severe psychiatric disorders with lower prevalence rates or emergence at later ages. The differential genetic correlations with common and relatively rare disorders suggests a continuum of genetic effects varying from very specific variants, variants which underlie either common or less common disorders, to variants which underlie most psychiatric problems. The presence of these universal variants is supported by genetic correlations between common and less common disorders, such as ADHD and schizophrenia.[2, 63] The latter set of variants may be better detected with measures of global psychopathology in older children, when thought disorders such as schizophrenia and bipolar disorder occur.

A limitation of this study is the large heterogeneity in measures of psychopathology. On the one hand, this variety of methods is an advantage, since any associations detected are expected to be more generalizable. On the other hand, it might limit the detectability of less robustly associated variants. This lack of power to identify in this study more loci probably stems mostly from insufficient sample size, but also from measurement error and low numbers of participating children with high psychiatric problems, as all cohorts were population-based.

Finally, as in any other GWAS study, the extent to which the found associations can be interpreted causally is difficult. Due to linkage disequilibrium it is unclear whether the two top variants have causal influence on psychopathology or are a marker for other causal variants. The same is true for the association of *SBF2* with total psychiatric problems. However, the association of predicted *SBF2* gene products with neuroticism and psychiatric problems, as well as the influence on myelination in an experimental mouse model, suggest a causal role.

In conclusion, this GWAS of total psychiatric problem scores suggests that common genetic variants exist that are associated simultaneously with internalizing, externalizing, attention and other psychiatric problems in childhood. The pleiotropy was not restricted to psychiatric phenotypes, but also included intelligence, educational attainment, wellbeing, smoking, body fat and number of children in adulthood. Interestingly, we did not find shared genetic effects with autism, schizophrenia and bipolar disorder. Two novel loci were genome-wide significant, though, the low sample size and modest imputation quality necessitate replication before firm conclusions can be drawn whether they influence total psychiatric problems. Furthermore, we found evidence that the gene *SBF2*, which was previously known to be associated with neuroticism, is also implicated in general psychopathology in children. Our results merit further investigation for confirmation and exploration of potential causal mechanisms.

## Data Availability

For summary statistics please contact Generation R data management and the corresponding author. Summary statistics of the meta-analysis will be made public after publication.

## 1958 British Birth Cohort

This work made use of data and samples generated by the 1958 Birth Cohort (NCDS), which is managed by the Centre for Longitudinal Studies at the UCL Institute of Education, funded by the Economic and Social Research Council (grant number ES/M001660/1). Access to these resources was enabled via the 58READIE Project funded by Wellcome Trust and Medical Research Council (grant numbers WT095219MA and G1001799). A full list of the financial, institutional and personal contributions to the development of the 1958 Birth Cohort Biomedical resource is available at http://www2.le.ac.uk/projects/birthcohort/1958bc/about/ contributors-funders. The Medical Research Council funded the 2002–2004 clinical follow-up of the 1958 birth cohort (grant G0000934). Genotyping was undertaken as part of the Wellcome Trust Case-Control Consortium (WTCCC) under Wellcome Trust award 076113, and a full list of the investigators who contributed to the generation of the data is available at www.wtccc.org.uk. This research used resources provided by the Type 1 Diabetes Genetics Consortium, a collaborative clinical study sponsored by the National Institute of Diabetes and Digestive and Kidney Diseases (NIDDK), National Institute of Allergy and Infectious diseases, National Human Genome Research Institute, National Institute of Child Health and Human Development, and Juvenile Diabetes Research Foundation International (JDRF) and supported by U01DK062418. The research was supported by the National Institute for Health Research Biomedical Research Centre at Great Ormond Street Hospital for Children NHS Foundation Trust and University College London.

## ALSPAC

We are extremely grateful to all the families who took part in this study, the midwives for their help in recruiting them, and the whole ALSPAC team, which includes interviewers, computer and laboratory technicians, clerical workers, research scientists, volunteers, managers, receptionists and nurses. The UK Medical Research Council and Wellcome (Grant ref: 102215/2/13/2) and the University of Bristol provide core support for ALSPAC. This publication is the work of the authors and they will serve as guarantors for the contents of this paper. GWAS data was generated by Sample Logistics and Genotyping Facilities at Wellcome Sanger Institute and LabCorp (Laboratory Corporation of America) using support from 23andMe.

## BREATHE

The research leading to these results has received funding from the European Research Council under the ERC Grant Agreement number 268479 - the BREATHE project. We are acknowledged with all the children and their families participating into the BREATHE project for their altruism.

SA and NVT are funded by Juan de la Cierva Programme (Ministry of Science, Innovation and Universities - Spanish State Research Agency, ref. IJCI-2017-34068, and ref. FJC-2018-038085-I).

## CADD

AGW is supported by NIAAA grant 3T32AA7464-38 S1 (PI:Paula Hoffman); Data collection and genotyping funded by DA05131,DA011015, & DA012845

## CATSS

The CATSS is supported by the Swedish Council for Working Life and Social Research, the Swedish Research Council, Systembolaget, the National Board of Forensic Medicine, the Swedish Prison and Probation Service, Bank of Sweden Tercentenary Foundation, the Söderström-Königska foundation, and the Karolinska Institutet Center of Neurodevelopmental Disorders (KIND).

## COPSAC

We express our deepest gratitude to the children and families of the COPSAC cohort study for all their support and commitment. We acknowledge and appreciate the unique efforts of the COPSAC research team. All funding received by COPSAC is listed on www.copsac.com. The Lundbeck Foundation (Grant no R16-A1694); The Ministry of Health (Grant no 903516)l; Danish Council for Strategic Research (Grant no 0603-00280B) and The Capital Region Research Foundation have provided core support to the COPSAC research center.

## FiNNTWIN12

FinnTwin12 Study is conducted by the University of Helsinki in close collaboration with Indiana University. Data collection was funded by the National Institute of Alcohol Abuse and Alcoholism and by the Academy of Finland. The present analysis was supported by the Academy of Finland (grants 265240 & 312073)

## GenR

The Generation R Study is conducted by the Erasmus Medical Center in close collaboration with the Erasmus University Rotterdam, Faculty of Social Sciences, the Municipal Health Service Rotterdam area, and the Stichting Trombosedienst and Artsenlaboratorium Rijnmond (STAR), Rotterdam. We gratefully acknowledge the contribution of general practitioners, hospitals, midwives and pharmacies in Rotterdam.

The Generation R Study is made possible by financial support from: Erasmus Medical Center, Rotterdam, and the Netherlands Organization for Health Research and Development (ZonMw). A. Neumann and H. Tiemeier are supported by a grant of the Dutch Ministry of Education, Culture, and Science and the Netherlands Organization for Scientific Research (NWO grant No. 024.001.003, Consortium on Individual Development). The work of H. Tiemeier is further supported by a European Union’s Horizon 2020 research and innovation program (Contract grant number: 633595, DynaHealth) and a NWO-VICI grant (NWO-ZonMW: 016.VICI.170.200). We would like to thank Anis Abuseiris, Karol Estrada, Dr. Tobias A. Knoch, and Rob de Graaf as well as their institutions Biophysical Genomics, Erasmus MC Rotterdam, The Netherlands, and especially the national German MediGRID and Services@MediGRID part of the German D-Grid, both funded by the German Bundesministerium fuer Forschung und Technology under grants #01 AK 803 A-H and # 01 IG 07015 G for access to their grid resources.

## Gini-Lisa

The authors thank all families for participation in the studies and the LISAplus and GINIplus study teams for their excellent work.

## Glaku

The study has been supported by Academy of Finland, University of Helsinki, Finnish Foundation for Pediatric Research, Sigrid Juselius Foundation, Jalmari and Rauha Ahokas Foundation, Signe and Ane Gyllenberg Foundation, Yrjo Jahnsson Foundation, Juho Vainio Foundation, Emil Aaltonen Foundation, and Ministry of Education and Culture, Finland. The 352 samples were genotyped at the Genotyping and Sequencing Core Facility of the Estonian Genome Centre, University of Tartu.

## INMA

The INMA project was funded by grants from Instituto de Salud Carlos III (Red INMA G03/176 and CB06/02/0041). The INMA-Sabadell cohort received funding from Instituto de Salud Carlos III (FIS-FEDER: PI041436 and PI081151), Generalitat de Catalunya-CIRIT 1999SGR 00241, and EU sixth framework project NEWGENERIS FP6-2003-Food-3-A-016320. The INMA-Valencia cohort received funding from UE (FP7-ENV-2011 cod 282957 and HEALTH.2010.2.4.5-1), and from Instituto de Salud Carlos III (FIS-FEDER: 03/1615, 04/1509, 04/1112, 04/1931, 05/1079, 05/1052, 06/1213, 07/0314, 09/02647, 11/0178, 11/01007, 11/02591, 11/02038, 13/1944, 13/2032, 14/00891, 14/01687). The authors also would particularly like to thank all the participants of INMA project for their generous collaboration.

NV-T is funded by a pre-doctoral grant from the Agència de Gestió d’Ajuts Universitaris i de Recerca (2017 FI_B 00636) Generalitat de Catalunya – Fons Social Europeu

SA is supported by a Sara Borrell grant from the Instituto de Salud Carlos III (CD14/00214).

## MUSP

The authors thank MUSP participants, the MUSP Research Team, the MUSP data collection teams, the Mater Misericordiae Hospital and the Schools of Social Science, Public Health, and Medicine, at The University of Queensland for their support and, the National Health and Medical Research Council (NHMRC).

## NFBC1986

Sigrid Juselius Foundation and the Strategic Research Funding from the University of Oulu. NFBC1966 and 1986 have received financial support from the Academy of Finland (project grants 104781, 120315, 129269, 1114194, 24300796, Center of Excellence in Complex Disease Genetics and SALVE), University Hospital Oulu, Biocenter, University of Oulu, Finland (75617), NIHM (MH063706, Smalley and Jarvelin), Juselius Foundation, NHLBI grant 5R01HL087679-02 through the STAMPEED program (1RL1MH083268-01), NIH/NIMH (5R01MH63706:02), the European Commission (EURO-BLCS, Framework 5 award QLG1-CT-2000-01643), ENGAGE project and grant agreement HEALTH-F4-2007-201413, EU FP7 EurHEALTHAgeing −277849, the Medical Research Council, UK (G0500539, G0600705, G1002319, PrevMetSyn/SALVE) and the MRC, Centenary Early Career Award. The program is currently being funded by the H2020 DynaHEALTH action (grant agreement 633595) and academy of Finland EGEA-project (285547). The DNA extractions, sample quality controls, biobank up-keeping and aliquotting was performed in the National Public Health Institute, Biomedicum Helsinki, Finland and supported financially by the Academy of Finland and Biocentrum Helsinki. We thank the late Professor Paula Rantakallio (launch of NFBCs), and Ms Outi Tornwall and Ms Minttu Jussila (DNA biobanking). The authors would like to acknowledge the contribution of the late Academian of Science Leena Peltonen.

## NTR

NTR warmly thanks all participating twin families. We acknowledge funding from multiple sources, including The Netherlands Organization for Scientific Research (NWO 480-15-001/674): Netherlands Twin Registry Repository: researching the interplay between genome and environment Consortium on Individual Development (CID NWO grant number 024.001.003), Genetic influences on stability and change in psychopathology from childhood to young adulthood (ZonMW 912-10-020), Biobanking and Biomolecular Resources Research Infrastructure (BBMRI –NL, 184.021.007); the Avera Institute, Sioux Falls, South Dakota (USA) and Grand Opportunity grants 1RC2 MH089951 and 1RC2 MH089995).

## RAINE

This study was supported by the National Health and Medical Research Council of Australia [grant numbers 572613 and 40398] and the Canadian Institutes of Health Research [grant number MOP-82893]. The authors are grateful to the Raine Study participants and their families, and to the Raine Study research staff for cohort coordination and data collection. The authors gratefully acknowledge the NH&MRC for their long term funding to the study over the last 25 years and also the following institutes for providing funding for Core Management of the Raine Study: The University of Western Australia (UWA), Curtin University, the Raine Medical Research Foundation, the UWA Faculty of Medicine, Dentistry and Health Sciences, the Telethon Kids Institute, the Women’s and Infant’s Research Foundation (King Edward Memorial Hospital), Murdoch University, The University of Notre Dame (Australia), and Edith Cowan University. The authors gratefully acknowledge the assistance of the Western Australian DNA Bank (National Health and Medical Reserach Council of Australia National Enabling Facility). Andrew Whitehouse is funded by Senior Research Fellowship from the NHMRC (1077966). We would also like to acknowledge the Raine Study participants for their ongoing participation in the study, and the Raine Study Team for study co-ordination and data collection. This work was supported by resources provided by the Pawsey Supercomputing Centre with funding from the Australian Government and Government of Western Australia.

## TCHAD

The TCHAD study is funded by the Swedish Council for Working Life and Social Research (project 2004-0383) and the Swedish Research Council (2004-1415).

## TEDS

We gratefully acknowledge the ongoing contribution of the participants in TEDS and their families. TEDS is supported by UK Medical Research Council Program Grant MR/M021475/1 (and previously Grant G0901245) (to R.P.), with additional support from National Institutes of Health Grant AG046938. The research leading to these results has also received funding from the European Research Council under the European Union’s Seventh Framework Programme (FP7/2007-2013)ERC Grant Agreement 295366. R.P. is supported by Medical Research Council Professorship Award G19/2. KR is supported by the Sir Henry Wellcome Postdoctoral fellowship.

## TRAILS

TRAILS (TRacking Adolescents’ Individual Lives Survey) is a collaborative project involving various departments of the University Medical Center and University of Groningen, the University of Utrecht, the Radboud Medical Center Nijmegen, and the Parnassia Group, all in the Netherlands. TRAILS has been financially supported by various grants from the Netherlands Organization for Scientific Research NWO (Medical Research Council program grant GB-MW 940-38-011; ZonMW Brainpower grant 100-001-004; ZonMw Risk Behavior and Dependence grants 60-60600-97-118; ZonMw Culture and Health grant 261-98-710; Social Sciences Council medium-sized investment grants GB-MaGW 480-01-006 and GB-MaGW 480-07-001; Social Sciences Council project grants; GB-MaGW 452-04-314 and GB-MaGW 452-06-004; NWO large-sized investment grant 175.010.2003.005; NWO Longitudinal Survey and Panel Funding 481-08-013 and 481-11-001; NWO Vici 016.130.002 and 453-16-007/2735; NWO Gravitation 024.001.003), the Dutch Ministry of Justice (WODC), the European Science Foundation (EuroSTRESS project FP-006), the European Research Council (ERC-2017-STG-757364 en ERC-CoG-2015-681466), Biobanking and Biomolecular Resources Research Infrastructure BBMRI-NL (CP 32), the Gratama foundation, the Jan Dekker foundation, the participating universities, and Accare Centre for Child and Adolescent Psychiatry. Statistical analyses were carried out on the Genetic Cluster Computer (http://www.geneticcluster.org), which is financially supported by the Netherlands Scientific Organization (NWO 480-05-003) along with a supplement from the Dutch Brain Foundation. We are grateful to everyone who participated in this research or worked on this project to make it possible.

## YFS

YFS has been financially supported by the Academy of Finland: grants 286284, 134309 (Eye), 126925, 121584, 124282, 129378 (Salve), 117787 (Gendi), and 41071 (Skidi); the Social Insurance Institution of Finland; Competitive State Research Financing of the Expert Responsibility area of Kuopio, Tampere and Turku University Hospitals (grant X51001); Juho Vainio Foundation; Paavo Nurmi Foundation; Finnish Foundation for Cardiovascular Research; Finnish Cultural Foundation; The Sigrid Juselius Foundation; Tampere Tuberculosis Foundation; Emil Aaltonen Foundation; Yrjö Jahnsson Foundation; Signe and Ane Gyllenberg Foundation; Diabetes Research Foundation of Finnish Diabetes Association; and EU Horizon 2020 (grant 755320 for TAXINOMISIS); European Research Council (grant 742927 for MULTIEPIGEN project); and The Wihuri Foundation. We thank the teams that collected data at all measurement time points; the persons who participated as both children and adults in these longitudinal studies; and biostatisticians Irina Lisinen, Johanna Ikonen, Noora Kartiosuo, Ville Aalto, and Jarno Kankaanranta for data management and statistical advice.

## Supplemental

Figure S1: Quality control steps of individual cohort results

Figure S2: Forest plot of results for rs10767094

Figure S3: Forest plot of results for rs202005905

Figure S4: Forest plot of results for rs72854494

Figure S5: Quantile-quantile plot of observed -log 10 p values vs expected -log 10 p values assuming chance findings in gene based analysis. Diagonal line indicates a p value distribution compatible with chance finding. Upward deviations indicate p values more significant than expected.

Figure S6: Manhattan plot of -log 10 p values vs SNP position for gene based analysis. Genes above the red horizontal line indicate genome-wide significant findings.

## Supplement

Table S1: Genes with genome-wide suggestive (p<3e-4) results

Table S2: Tissue expression analysis (neural tissues)

Table S3: Tissue expression analysis (organs)

